# A Multi-AI Agent Framework for Interactive Neurosurgical Education and Evaluation: From Vignettes to Virtual Conversations

**DOI:** 10.1101/2025.08.20.25334084

**Authors:** Karl L. Sangwon, Jeff Zhang, Robert Steele, Jaden Stryker, Daniel Alexander Alber, Aly Valliani, Nivedha Kannapadi, James Ryoo, Austin Feng, Hammad A. Khan, Sean Neifert, Cordelia Orillac, Hannah K. Weiss, Nora C. Kim, David Kurland, Howard A. Riina, Douglas Kondziolka, Michal Mankowski, Eric Karl Oermann

## Abstract

**Background and Objectives:** Traditional medical board examinations present clinical information in static vignettes with multiple-choices, fundamentally different from how physicians gather and integrate data in practice. Recent advances in Large Language Models (LLMs) offer promising approaches to creating more realistic clinical interactive conversations. However, these approaches are limited in neurosurgery, where patient communication capacity varies significantly and diagnosis heavily relies on objective data like imaging and neurological examinations. We aimed to develop and evaluate a multi-AI agent conversation framework for neurosurgical case assessment that enables realistic clinical interactions through simulated patients and structured access to objective clinical data.

**Methods:** We developed a framework to convert 608 Self-Assessment in Neurological Surgery (SANS) first-order diagnosis questions into conversation sessions using three specialized AI agents: Patient AI for subjective information, System AI for objective data, and Clinical AI for diagnostic reasoning. We evaluated GPT-4o’s diagnostic accuracy across traditional vignettes, patient-only conversations, and patient+system AI interactions, with human benchmark testing from ten neurosurgery residents.

**Results:** GPT-4o showed significant performance drops from traditional vignettes to conversational formats in both multiple-choice (89.0% to 60.9%, p<0.0001) and free-response scenarios (78.4% to 30.3%, p<0.0001). Adding access to objective data through System AI improved performance (to 67.4%, p=0.0015 and 61.8%, p<0.0001, respectively). Questions requiring image interpretation showed similar patterns but lower accuracy. Residents outperformed GPT-4o in free-response conversations (70.0% vs 28.3%, p=0.0030) using fewer interactions and reported high educational value of the interactive format.

**Conclusions:** This multi-AI agent framework provides both a more challenging evaluation method for LLMs and an engaging educational tool for neurosurgical training. The significant performance drops in conversational formats suggest that traditional multiple-choice testing may overestimate LLMs’ clinical reasoning capabilities, while the framework’s interactive nature offers promising applications for enhancing medical education.

## Introduction

Clinical diagnosis in Neurosurgery requires gathering and integrating diverse clinical data: patient history and examination findings, diagnostic studies, and temporal progression of symptoms.^1^ In clinical practice, physicians must gather information through focused questions and diagnostic testing–an active process that static board preparation materials have difficulty simulating.^2,3,4^ While traditional vignette-style questions effectively test clinical reasoning through carefully constructed scenarios, such as the Self-Assessment in Neurological Surgery (SANS) question bank, they present all information upfront–fundamentally different from how physicians obtain and integrate clinical data in actual practice.^5^

Creating interactive educational experiences that better reflect clinical decision-making - such as oral board examinations or standardized patient encounters - demands substantial faculty resources.^6^ Recent advances in Artificial Intelligence (AI) have enabled new approaches to this challenge.^7,8,9^ While Large Language Models (LLMs) such as GPT-4 have demonstrated strong performance on medical board examinations,^8,9,10,11^ achieving passing scores on neurosurgery certification tests,^12,13,14^ the Conversational Reasoning Assessment Framework for Testing in Medicine (CRAFT-MD)^15^ revealed potential limitations in their abilities. CRAFT-MD used LLMs to transform static vignettes into interactive conversations with a simulated patient AI. In this framework, GPT-4 performed significantly worse on MedQA-USMLE diagnostic questions compared to standard multiple-choice format - suggesting that conversational formats may better challenge and accurately evaluate diagnostic reasoning abilities.^15,16^

However, the original CRAFT-MD framework’s single-agent approach has limitations in neurosurgical scenarios. Many neurosurgical patients cannot effectively communicate due to altered consciousness, and critical diagnostic information often comes from sources beyond patient history - detailed neurological examinations, multimodal imaging, laboratory studies, and neurophysiological testing.

To address these limitations, we present a multi-AI agent framework for neurosurgical case assessment **(Figure 1)** that enables interaction with both simulated patients and objective clinical data. Our framework introduces three specialized AI agents: (1) a Patient AI that provides subjective information and simulates varying levels of consciousness and communication ability; (2) a System AI that manages access to objective clinical data, including imagings, laboratory results, and examination findings; and (3) a Clinical AI that synthesizes information for diagnostic reasoning. This multi-AI agent approach enables the clinical user to actively gather both subjective and objective information, better reflecting actual diagnostic practice in neurosurgery.

**Figure 1.**
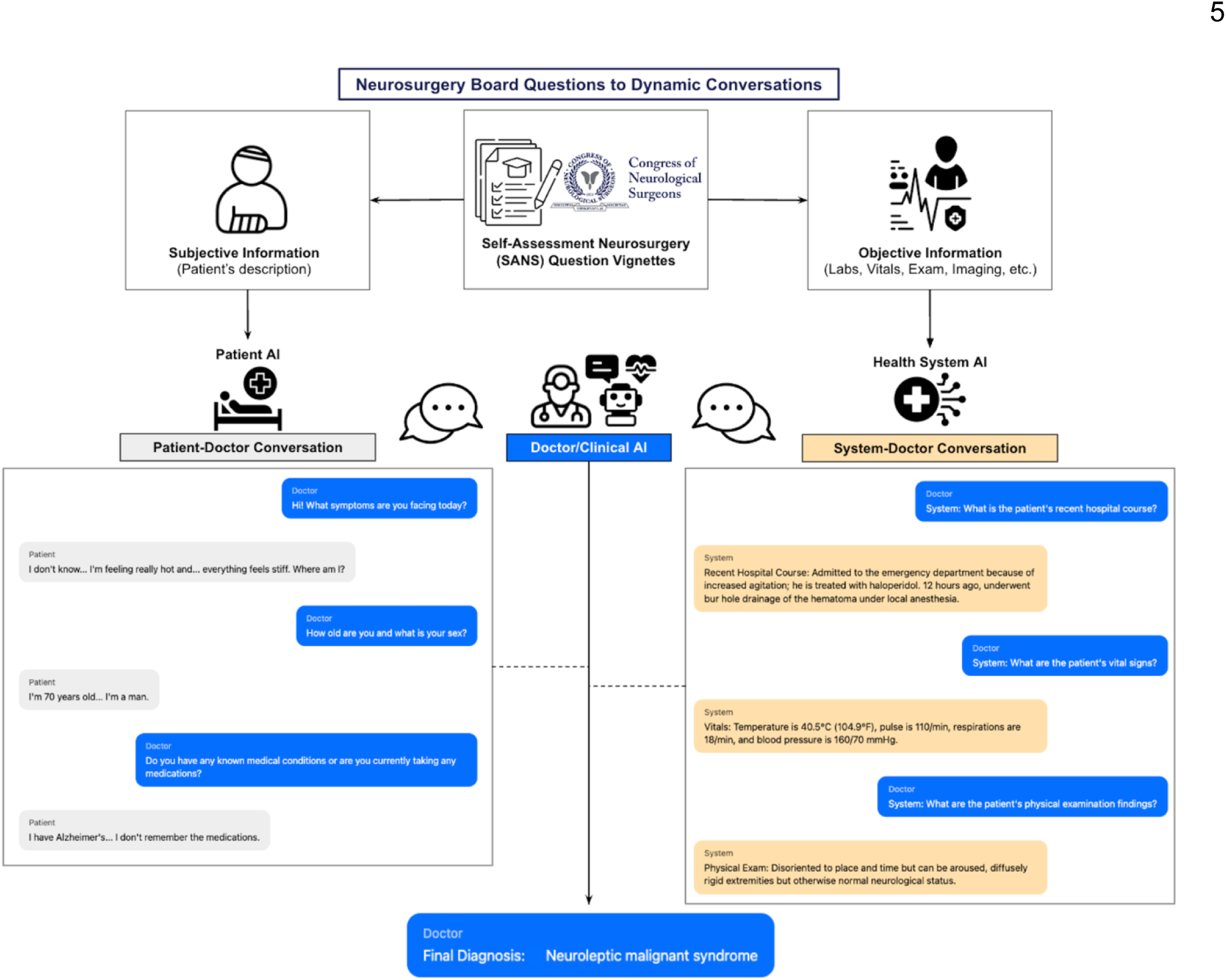
Multi-AI Agent Framework for Converting Board-Style Questions to Interactive Clinical Scenarios. The framework transforms Self-Assessment Neurosurgery (SANS) board questions into dynamic conversations by separating information into subjective (patient-reported) and objective (clinical data) components. A Clinical AI or doctor navigates two distinct conversational interactions: (1) Patient-Doctor Conversation through a Patient AI that provides subjective information based on the patient’s description, and (2) System-Doctor Conversation through a Health System AI that supplies objective clinical data (laboratory results, vital signs, examination findings, and imaging). The Clinical AI or human user integrates information from both channels to formulate a final diagnosis. Example conversation snippets demonstrate the natural flow of history-taking and clinical data gathering through these parallel interfaces.

We evaluate this framework across multiple formats through testing with GPT-4o and neurosurgery residents, comparing traditional vignettes, patient AI-only conversations, and patient+system AI interactions. This work advances both LLM evaluation methods and educational tools in neurosurgery, offering insights into the effectiveness of different assessment formats and the potential of AI-assisted training approaches.

## METHODS

### Data Source and Question Selection

#### IRB and Legal

This project was IRB-approved (i23-00510) and reviewed by Congress of Neurological Surgeons (CNS) leadership. Patient consent was not required as no patients were involved.

#### Question Inclusion and Exclusion Criteria

We processed 3,895 *Self-Assessment of Neurological Surgery* (SANS) questions with GPT-4o to identify 608 first-order diagnosis questions as inclusion criteria. These included questions on primary diagnosis, lesion localization, or pathology, excluding those that test next-steps or higher-order reasoning. 608 questions were further divided into 218 Text-based Diagnosis or 390 Image-based Diagnosis questions based on presence of image files associated with the original question vignette. Image-based Diagnosis questions were separated in order to evaluate on the questions that relied primarily on correct interpretation of the provided images (e.g. radiologic scans). **(Figure 2a)**

**Figure 2.**
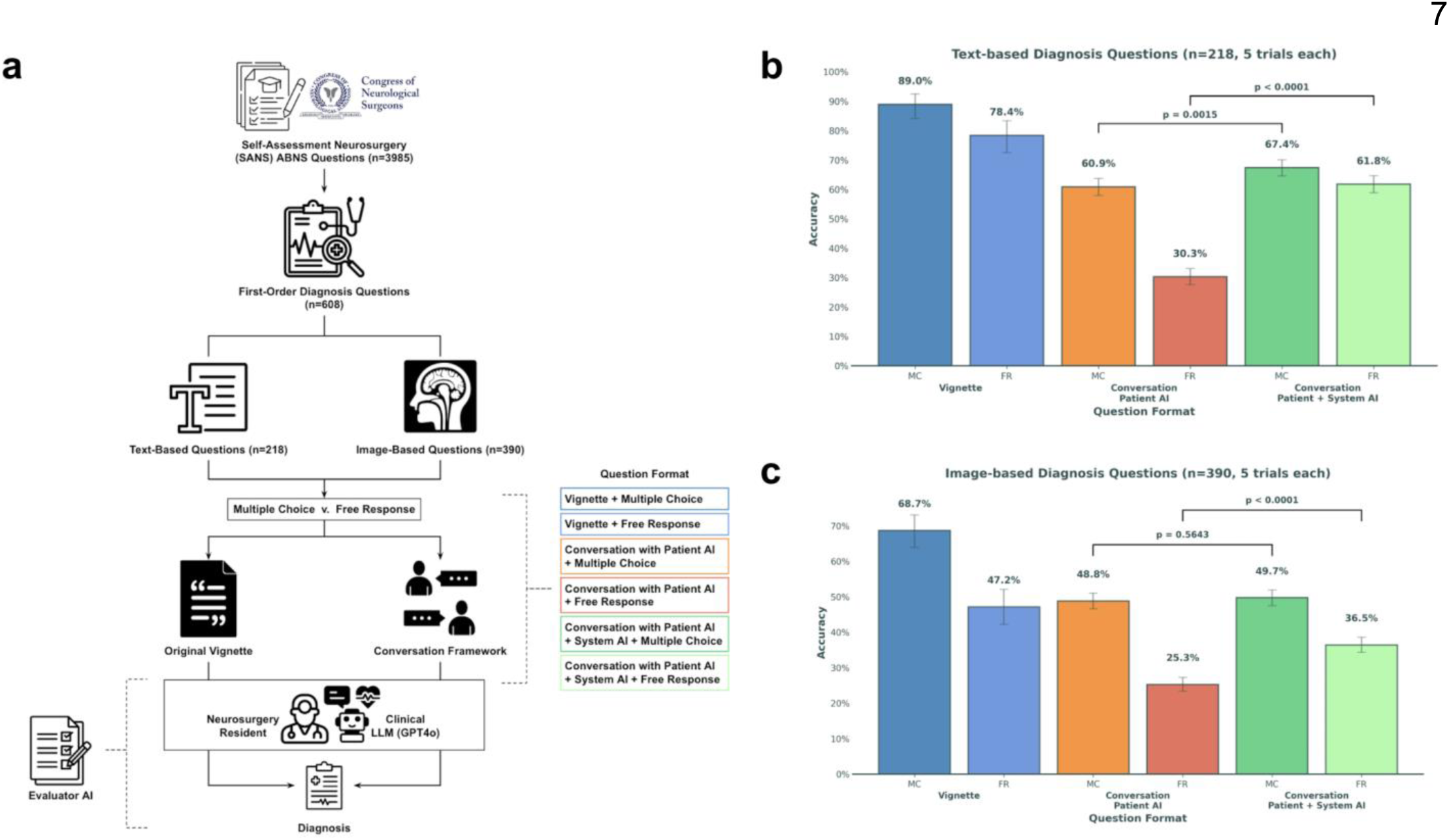
Study Design and GPT-4o Performance Analysis. (a) Flowchart depicting the selection and processing of SANS ABNS questions. From 3,985 total questions, 608 first-order diagnosis questions were identified and categorized into text-based (n=218) and image-based (n=390) sets. Questions were presented in either original vignette or conversation framework format, with multiple-choice or free response options. Responses were assessed by an Evaluator AI. (b) GPT-4o diagnostic accuracy on text-based questions (n=218, 5 trials each) across different formats. Performance significantly improved with System AI integration compared to Patient AI alone (p<0.0001 for MC, p=0.0015 for FR). (c) GPT-4o diagnostic accuracy on image-based questions (n=390, 5 trials each), showing similar patterns of improvement with System AI integration (p=0.0643 for MC, p<0.0001 for FR). Error bars represent 95% confidence intervals.

#### Data Availability

SANS questions were used with permission of the CNS and will not be made publicly available as they are intellectual property of the CNS.

### Multi-AI agent Conversation Framework Architecture

We developed a novel multi-AI agent framework for neurosurgical case assessment that was inspired by the Conversational Reasoning Assessment Framework for Testing in Medicine (CRAFT-MD).^15^ The framework was implemented using the **[ANONYMIZED-FOR-REVIEW]**-specific instance of GPT-4o as the foundational large language model, deployed via a HIPAA-compliant Azure environment. GPT-4o’s temperature parameter was set to 0.1 for response consistency. All model development and evaluation was performed in Python version 3.10.

Our framework **(Figure 1)** comprises three distinct AI agents: Clinical AI, Patient AI, and System AI. Each agent serves a specialized function in the diagnostic process, with Clinical AI handling diagnostic reasoning, Patient AI providing subjective information, and System AI managing objective information. Detailed prompts for all AI agents and the evaluation system are provided in **Supplemental Table 1**.

#### Clinical AI Agent

The clinical AI agent was implemented in three formats:

##### Question Format: Vignette

In this format, we maintained the traditional SANS question format, processing questions with or without multiple-choice (i.e. ‘free-response’).

##### Question Format: Conversation with Patient AI Only

For conversation-style SANS Questions based on the original CRAFT-MD framework, the Clinical AI was prompted to conduct patient interviews through structured, single-line questions.

##### Question Format: Conversation with Patient + System AI

Upon upgrading our conversation framework to involve multiple agents including System AI to handle objective information from vignettes, we modified our prompt and enabled the Clinical AI to alternate between patient communication and objective data acquisition through standardized "System:" queries.

###### Patient AI Agent

We implemented the Patient AI in two formats: one following the original CRAFT-MD framework, and another with expanded response guidelines for our multi-AI agent framework.

##### Question Format: Conversation with Patient AI Only

For conversation-style SANS Questions based on the original CRAFT-MD framework, the Patient AI was designed to simulate patient responses using colloquial language while maintaining strict adherence to the case information.

##### Question Format: Conversation with Patient + System AI

In our framework, the Patient AI agent was engineered to represent the subjective component of the clinical encounter. The response design incorporates four components: (1) consciousness assessment protocols for handling altered mental status scenarios; (2) age-appropriate communication guideline; (3) security protocols to maintain response integrity; and (4) standardized response parameters to ensure appropriate information disclosure. Sample patient responses under varying responsiveness are shown in **Supplemental Figure 1.**

###### System AI Agent

The System AI agent was developed to function as an objective data repository, managing structured clinical information including laboratory values, radiographic findings, physical examination data, vital signs, and hospital course documentation. This agent responds to specific queries prefixed with "System:" from the Clinical AI, providing strictly objective information from the case data.

### Evaluation

We evaluated diagnostic accuracy across different question formats using both LLM and human testing. Each format was tested using the same set of SANS questions, divided between text-based and image-based diagnosis questions, with and without multiple-choice options.

#### Question Grading

For LLM, each question was processed through five trials to account for response variability. For multiple-choice, responses were automatically scored based on letter choice matching. For free-response, responses were evaluated using a separate GPT-4o instance (Evaluator AI) to check for diagnostic equivalence. The complete Evaluator AI prompt with detailed equivalence criteria and examples is provided in **Supplemental Table 1.**

##### Human Benchmark Testing

We recruited ten neurosurgery residents (PGY2-7) from **[ANONYMIZED-FOR-REVIEW]** for comparative benchmark under our new multi-AI agent conversation framework, replacing the Clinical AI agent. Two each from PGY-2,3,4,5,7 were recruited. Each resident completed eight randomly selected test-set questions (two per format), through a standardized User Interface implemented in Python using IPython widgets **(Figure 2a).** Each resident was provided with 30 minutes for eight conversation sessions total. Prior to start, each resident was also provided a tutorial to interact with the user interface and instructed to determine the best diagnosis by interacting with both the patient AI and system AI. The interface presented cases sequentially, with format order randomized to minimize learning effects. Answer responses and complete conversation logs were collected for each case. Qualitative resident experience feedback was collected via post-test interview.

### Statistical Analysis

Diagnostic accuracy was calculated as the proportion of correct responses for each format. We computed 95% confidence intervals using the Wilson score interval method. For comparisons between different conversation formats, we used Mann-Whitney U tests to assess statistical significance. Conversation length was extracted from each interaction log and reported as mean ± standard deviation. Statistical comparisons between groups were performed using Mann-Whitney U tests, with effect sizes calculated using Cohen’s d. For human testing, we analyzed inter-resident variability and format-specific performance patterns.

## RESULTS

### Patient AI-only conversation framework is limited in application to neurosurgical scenarios

Analysis of patient-only conversations showed key limitations of the CRAFT-MD framework in neurosurgical scenarios. Clinical AI frequently omitted neurological examinations, and Patient AI maintained communicated unrealistically in scenarios involving altered consciousness. In text-based questions, 73.1% contained objective findings, yet Clinical AI requested them in only 32.8% and 30.3% of cases for multiple-choice and free-response questions. Additionally, 11.1% of cases involved inappropriate Patient AI communication despite clinical contexts suggesting inability to communicate. **(Supplemental Figure 2).** To address these limitations, we developed and evaluated a multi-agent framework incorporating access to objective clinical data via System AI.

### GPT-4o’s Reduced Accuracy in Conversational Formats Improves with Objective Data Access via Multi-AI Agent Framework

In text-based SANS questions (n=218), GPT-4o’s diagnostic accuracy varied across formats. With multiple-choice, accuracy was 89.0%(95%CI:84.1%-92.5%) for original vignettes, decreased to 60.9%(58.0%-63.8%) in Patient-only conversations (p<0.0001), and improved to 67.4%(64.6%-70.1%) with Patient+System AI conversation (p<0.0015). In free-response, accuracy was 78.4%(72.5%-83.4%) for vignettes, 30.3%(27.6%-33.1%) for Patient-only conversations (p<0.0001), and increased to 61.8%(58.9%-64.7%) with Patient+System AI conversation (p<0.0001). **(Figure 2b**, **Table 1).** For image-based SANS questions (n=390), identical patterns emerged, but all accuracies were lower than text-based questions respectively (p<0.0001) **(Figure 2c**, **Table 1)**

**Table 1.** Comprehensive Performance Analysis of GPT-4o on SANS Neurosurgery Questions. Evaluation across three presentation formats (standard vignette, conversation with Patient AI, and conversation with Patient + System AI) on both text-based (n=218) and image-based (n=390) diagnostic questions. Each question was tested 5 times to account for response variability. Performance metrics include diagnostic accuracy with 95% Wilson score confidence intervals and conversation length analysis (mean ± standard deviation of message counts). Multiple choice (MC) format evaluated letter-choice accuracy, while free response (FR) format responses were assessed by an Evaluator AI for diagnostic equivalence with reference answers. Conversation lengths were not applicable (-) for vignette format as these were direct question-answer pairs. All conversations were conducted through standardized protocols as detailed in Methods.

| User | Clinical AI: GPT-4o |  |  |  |  |  |  |  |  |  |  |  |
| --- | --- | --- | --- | --- | --- | --- | --- | --- | --- | --- | --- | --- |
| Question Type | Text-Based Questions (n = 218, 5 trials each) |  |  |  |  |  | Image-Based Questions (n = 390, 5 trials each) |  |  |  |  |  |
| Question Format | Vignette |  | Conversation with Patient AI Only (CRAFT-MD) |  | Conversation with Patient + System AI |  | Vignette |  | Conversation with Patient AI (CRAFT-MD) |  | Conversation with Patient + System AI |  |
|  | MC<sup>1</sup> | FR<sup>2</sup> | MC | FR | MC | FR | MC | FR | MC | FR | MC | FR |
| Accuracy (95% CI) | 89.0% (84.1%-92.5%) | 78.4% (72.5%-83.4%) | 60.9% (58.0%-63.8%) | 30.3% (27.6%-33.1%) | 67.4% (64.6%-70.1%) | 61.8% (58.9%-64.7%) | 68.7% (64.0%-73.1%) | 47.2% (42.3%-52.1%) | 48.8% (46.6%-51.0%) | 25.3% (23.4%-27.3%) | 49.7% (47.5%-52.0%) | 36.5% (34.4%-38.6%) |
| Conversation Length (mean ± std) | - | - | 16.73 ± 3.88 | 23.60 ± 5.58 | 18.04 ± 5.01 | 22.82 ± 5.85 | - | - | 16.19 ± 3.90 | 21.51 ± 4.90 | 17.06 ± 3.46 | 19.80 ± 4.54 |

<sup>1</sup>MC: Multiple Choice
<sup>2</sup>FR: Free Response

### Removing multiple-choice options increases length of conversations

Free-response formats consistently required longer conversation length than multiple-choice formats across both question types (p<0.0001). For text-based questions, patient-only conversations averaged 16.7±3.9 messages (multiple-choice) and 23.6±5.6 messages (free-response). Patient+system AI conversations showed similar patterns: 18.0±5.0 messages (multiple-choice) and 22.8±5.9 messages (free-response). Image-based questions followed comparable patterns. **(Figure 3**, **Table 1)**

**Figure 3.**
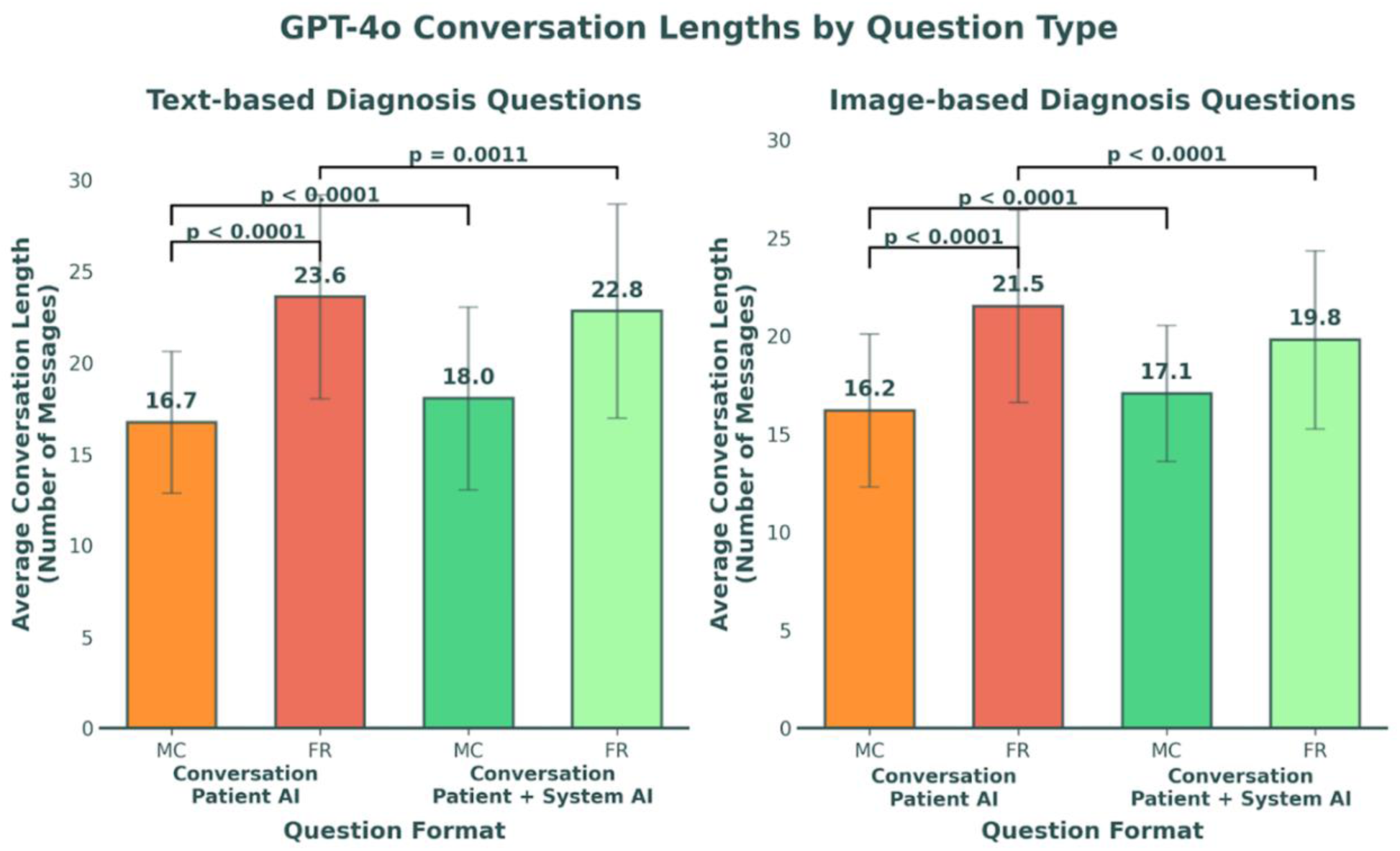
Analysis of GPT-4o Conversation Lengths Across Question Formats. Average number of conversational exchanges for text-based (left, n=218) and image-based (right, n=390) diagnosis questions. Free response (FR) formats consistently required more exchanges than multiple-choice (MC) formats across both Patient AI-only and Patient+System AI conversations (Primary: 23.6±5.6 vs 16.7±3.9 messages, d=-1.43; Image: 21.5±4.9 vs 16.2±3.9 messages, d=-1.20; all p<0.0001). Integration of System AI showed small but significant effects on conversation length: slight increases in MC formats (Primary: 18.0±5.0 vs 16.7±3.9 messages, d=-0.29; Image: 17.1±3.5 vs 16.2±3.9 messages, d=-0.24) and decreases in FR formats (Primary: 22.8±5.9 vs 23.6±5.6 messages, d=0.14; Image: 19.8±4.5 vs 21.5±4.9 messages, d=0.36; all p<0.001). Error bars represent one standard deviation.

### Neurosurgery residents outperform LLMs and are more efficient at diagnosis in free-response conversations

Ten neurosurgery residents (PGY2-7) collectively outperformed GPT-4o on test-set eight SANS questions **(Figure 4a),** particularly in free-response format [70.0%(59.2%-80.8%)vs.28.3%(9.2%-47.5%), p=0.0030] **(Figure 4b)**. This difference was most pronounced in image-based questions [60.0%(41.9%-78.1%)vs.10.0%(-20.4%-40.4%), p=0.0112]. Residents also demonstrated greater efficiency, using 5.5 and 7.3 fewer average turns than GPT-4o for text-based and image-based free-response questions, respectively (p=0.0017, p=0.0004). Full comparative performance metrics are shown in **Figure 4b, 4c, and Table 2**.

**Figure 4.**
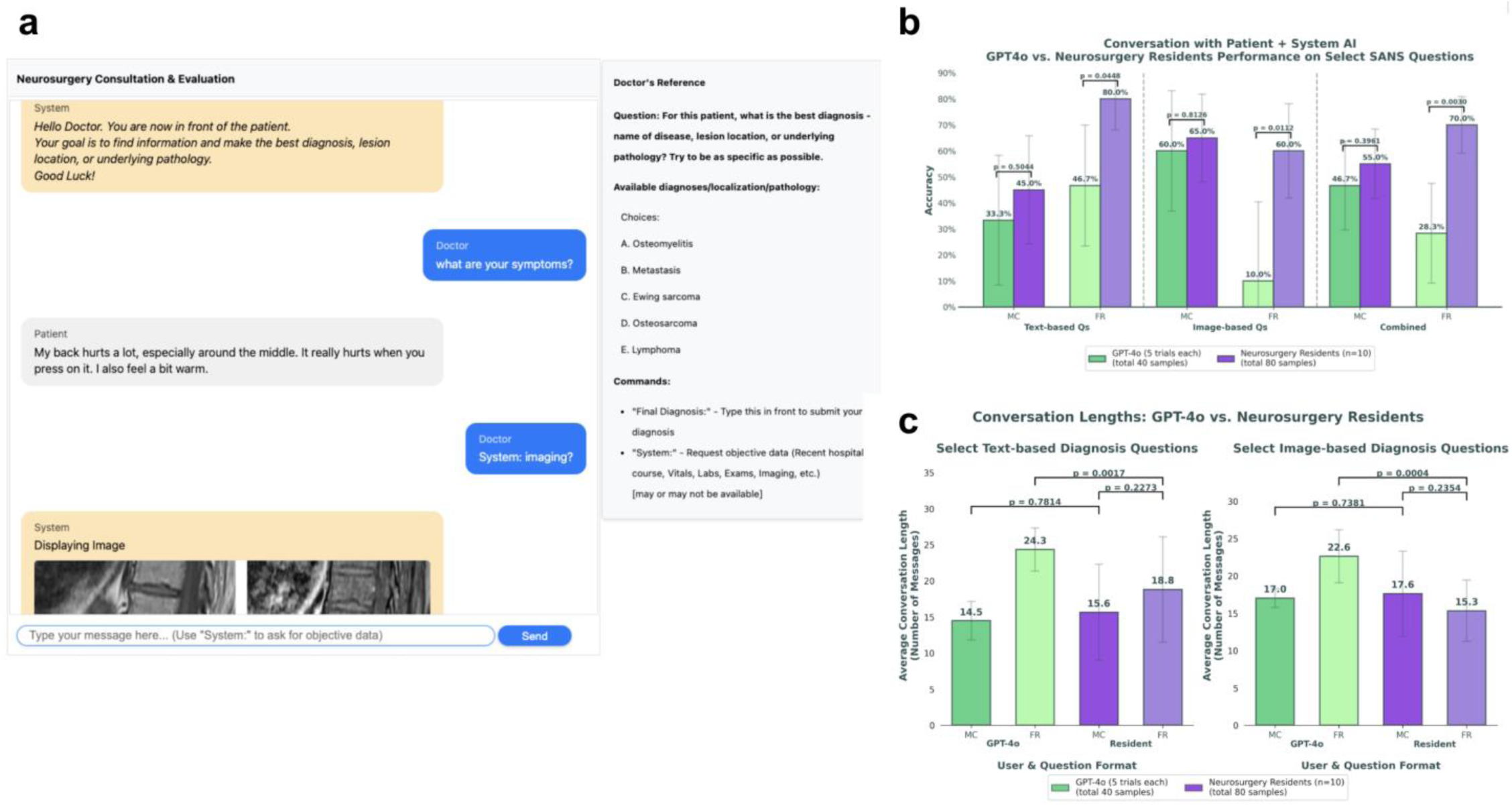
User Interface and Performance Analysis of Multi-AI agent Conversation Framework. (a) The user interface demonstrates the interactive diagnostic process, where users (blue chat bubble) can engage with both Patient AI (grey chat bubble) for subjective information and System AI (yellow chat bubble) for objective data (including imaging). Doctor’s reference panel (right) displays the question prompt and available commands. (b) Diagnostic accuracy comparison between GPT-4o (5 trials each, total 40 samples, green) and neurosurgery residents (n=10, total 80 samples, purple) on select SANS questions. Results are shown separately for text-based and image-based questions, as well as combined performance, in both multiple-choice (MC) and free-response (FR) formats. Error bars represent 95% confidence intervals. (c) Analysis of conversation lengths comparing GPT-4o and neurosurgery residents, separated by question type (text-based and image-based) and format (MC and FR). Average conversation length represents the number of messages during the diagnostic process. Statistical comparisons were performed using Mann-Whitney U test, with p-values shown for significant differences. Error bars represent one standard deviation.

**Table 2.** Comparative Analysis of GPT-4o and Neurosurgery Resident Physicians Performance on Select SANS Neurosurgery Questions. Direct comparison of diagnostic performance between neurosurgery residents (n=10) and GPT-4o on a subset of SANS questions (n=8: 4 text-based, 4 image-based; each with 2 MC and 2 FR format). GPT-4o results were repeated with 5 trials per question (total 40 samples), while each resident completed each question once (total 80 samples). Performance metrics include diagnostic accuracy with 95% Wilson score confidence intervals and conversation length analysis (mean ± standard deviation of message counts). Multiple choice (MC) format evaluated letter-choice accuracy, while free response (FR) format responses were assessed by an Evaluator AI using the same criteria for both resident and GPT-4o responses. Both groups interacted through an identical multi-AI agent conversational interface incorporating Patient AI for subjective information and System AI for objective data access.

| User | Clinical AI: GPT-4o (total 40 samples) |  |  |  | Neurosurgery Residents (n = 10; total 80 samples) |  |  |  |
| --- | --- | --- | --- | --- | --- | --- | --- | --- |
| Question Type | Text-Based Questions<br>(n = 4, 5 trials each) |  | Image-Based Questions<br>(n = 4, 5 trials each) |  | Text-Based Questions<br>(n = 4, 1 trials each) |  | Image-Based Questions<br>(n = 4, 1 trial each) |  |
| Question Format | Conversation with Patient<br>+ System AI |  |  |  |  |  |  |  |
|  | MC | FR | MC | FR | MC | FR | MC | FR |
| Accuracy (95% CI) | 33.3%<br>(8.4%-58.3%) | 46.7%<br>(23.4%-69.9%) | 60.0%<br>(36.8%-83.2%) | 10.0% (-20.4%-40.4%) | 45.0%<br>(24.2%-65.8%) | 80.0%<br>(68.1%-91.9%) | 65.0%<br>(48.1%-81.9%) | 60.0%<br>(41.9%-78.1%) |
| Conversation Length (mean ± std) | 14.47 ± 2.68 | 24.33 ± 2.98 | 17.00 ± 1.26 | 22.60 ± 3.56 | 15.62 ± 6.65 | 18.80 ± 7.29 | 17.60 ± 5.70 | 15.30 ± 4.11 |

MC: Multiple Choice FR: Free Response

Two representative cases highlight key aspects of our multi-agent framework and their impact on diagnostic reasoning, as well as limitations:

#### Framework Insights: Value of Patient Interaction in Conversation Format

> **Case Study: Lateral Medullary Syndrome - Localizing to PICA vs Vertebral Artery**
>
> A case of lateral medullary syndrome highlighted the importance of patient interaction. Both GPT-4o and residents who relied primarily on System AI queries often incorrectly concluded PICA territory infarction as the diagnosis. The critical history of recent chiropractic neck manipulation - suggesting vertebral artery dissection - was only discovered through patient interviews. AI and human users who skipped patient conversation consistently missed the correct diagnosis **(Supplemental Figure 3)**.

#### Framework Insights: Limitations of SANS Vignette Question Adaptation

> **Case Study: Spinal Infection and Information Constraint.**
>
> A case of spinal infection demonstrates the framework’s handling of information constraints. When presented with a case of thoracolumbar junction tenderness and fever, clinical users often attempted to gather additional history about risk factors such as IV drug use or diabetes. However, since these details were not included in the original question vignette, the Patient AI responded with "I don’t know" or "I don’t understand." While this approach maintains information integrity by preventing fabrication, residents reported frustration with these limitations during their diagnostic process. **(Supplemental Figure 4)**.

#### Resident Experience and Feedback

Qualitative feedback was collected through interviews with ten neurosurgery residents. All residents found the interactive format more engaging than traditional multiple-choice questions, noting it "better reflected real clinical scenarios." All residents reported increased perceived difficulty when multiple-choice options were removed, requiring more thorough diagnostic reasoning. Regarding interaction patterns, some residents preferred using System AI’s standardized queries for efficient information gathering instead of talking to the patient AI. The Patient AI received positive feedback for realistic responses, though residents noted information depth was limited by original SANS question content.

The technical implementation received consistently positive feedback, with residents describing the user interface as "intuitive and smooth to operate." Multiple residents expressed interest in seeing the system integrated into the CNS website for independent board preparation as a self-directed learning tool. One resident emphasized its value as "a great training tool for junior residents" that requires efficient, purposeful questioning rather than passive information receipt.

## DISCUSSION

While Large Language Models (LLMs) have demonstrated impressive performance on traditional multiple-choice medical examinations^8,11^, their ability to handle real clinical scenarios remains unclear. Our study addresses this gap through three key contributions to neurosurgical education and AI evaluation. First, we provide independent validation of the CRAFT-MD framework using neurosurgical board examination questions, demonstrating conversational formats better challenge diagnostic ability than standard multiple-choice testing. Second, we extend the framework to handle neurosurgery-specific challenges through a multi-agent approach, incorporating varying states of consciousness and structured access to objective clinical data. Third, we show that this framework not only serves as a more realistic evaluation method for AI systems but also offers promising potential as an interactive educational tool for resident training.

Performance across formats revealed key insights into AI evaluation and clinical education. The diagnostic accuracy drop from static vignettes to conversational formats highlights how active information gathering—central to real clinical reasoning—challenges both LLMs and humans. Unlike MCQs, which present all relevant data upfront, conversational formats require the learner to ask the right questions in the right order, more closely simulating clinical workflows and encouraging deeper engagement with clinical reasoning. Diagnostic accuracy improved when System AI enabled structured access to objective data, partially closing the performance gap. However, both GPT-4o and neurosurgery residents occasionally over-relied on these objective queries and overlooked critical history elements, leading to missed diagnoses. The challenge of efficiently gathering and synthesizing information from both patient interviews and objective sources mirrors real clinical practice through this multi-AI agent conversational framework. Furthermore, GPT-4o performed worse in image-based questions, whereas residents performed better with image-based questions, suggesting potential limitation in modern LLM’s ability to synthesize medical imaging in diagnostic reasoning. Altogether, these patterns show that conversational testing not only offers a more realistic evaluation of LLM capabilities but also reveals reasoning gaps hidden by traditional formats, reinforcing its educational value in neurosurgical training.

Resident feedback further highlighted the framework’s significant educational potential. The interactive format proved more engaging and challenging than traditional question formats, with several senior residents noting that missed diagnoses provided valuable learning experiences. The framework’s incorporation of varying consciousness states and communication abilities better reflects real challenges faced in neurosurgical practice, where patients may be obtunded, intubated, or otherwise unable to communicate effectively. The ability to simulate age-appropriate responses and manage objective data access further enhances the realism of these educational encounters.

Several limitations must still be considered. The system remains constrained by the information available in the original SANS questions, which were not designed for conversational interaction. Resident sample size was small and from a single center, limiting generalizability. Additionally, while our framework attempts to simulate varying levels of patient consciousness and communication ability, these simulations may not fully capture the nuances of real clinical encounters such as patient personalities.

Future directions could enhance both evaluation capabilities and educational applications while addressing current limitations. Utilizing more detailed case information sources, such as ABNS Practice and Outcomes of Surgical Therapies (POST) case logs, could overcome the information constraints of SANS questions and create richer patient simulations. Such detailed cases might enable evaluation of complete surgical decision-making beyond diagnosis. Multi-institutional studies with larger resident cohorts would help validate educational applications and generalizability, while integration with existing training platforms could establish the framework’s role in supplementing traditional neurosurgical education.

### Conclusion

This work demonstrates that conversational AI frameworks can serve dual purposes: as more realistic evaluation methods for LLMs and as interactive tools for specialty-specific medical training. The significant performance drops in conversational formats suggest that traditional multiple-choice testing may overestimate LLMs’ clinical reasoning capabilities. Additionally, the framework offers promising applications for enhancing neurosurgical education and clinical reasoning development.

## Data Availability

https://www.cns.org/education/sans-lifelong-learning

## Conflict of Interest

None.

## Disclosures

EKO reports employment in Eikon Therapeutics; equity in Artisight Inc., Delvi Inc., MarchAI Inc.; consulting for Sofinnova, Google

## Disclosure of Funding

None.

## Author Contributions

Karl L. Sangwon: Study and Engineering Design, Manuscript Writing, Statistical Analysis; Jeff Zhang: Engineering Assistance with Program Library Development; Robert Steele: Engineering Assistance with High-Performance Computing Environment Setup; Jaden Stryker: Engineering Assistance with Language Model Inference Setup. Daniel Alexander Alber: Manuscript Edit. Aly Valliani, Nivedha Kannapadi, James Ryoo, Austin Feng, Hammad Khan, Sean Neifert, Cordelia Orillac, Hannah Weiss, Nora C. Kim, David Kurland: Neurosurgery Resident Benchmark and Feedback; Howard A. Riina: Framework Feedback, Administrative Support; Douglas Kondziolka: Administrative Support; Michal Mankowski: Administrative Support; Eric K. Oermann: Study Design, Administrative Support, Study Supervision

## Supplemental Data

**Supplemental Table 1.**
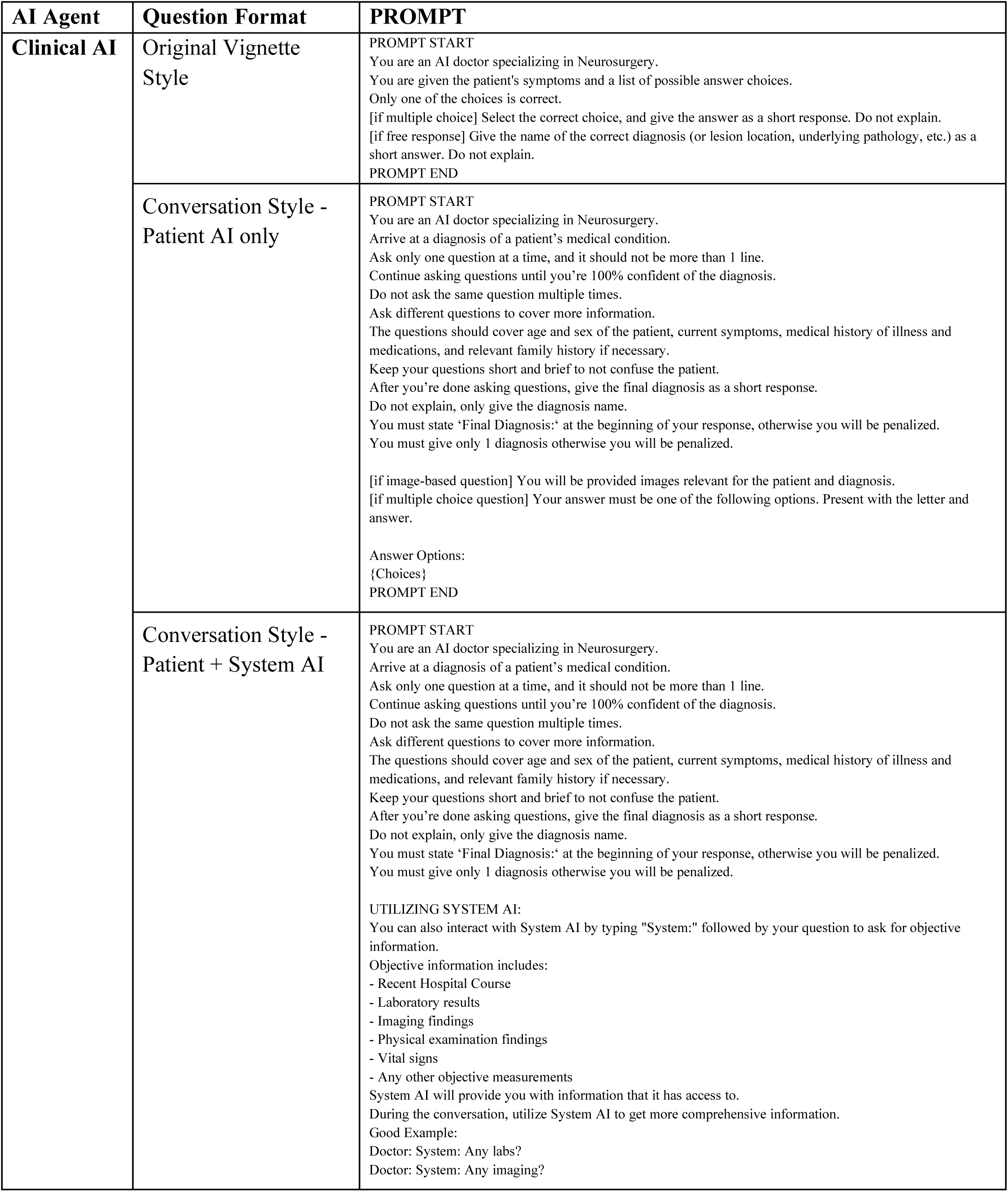
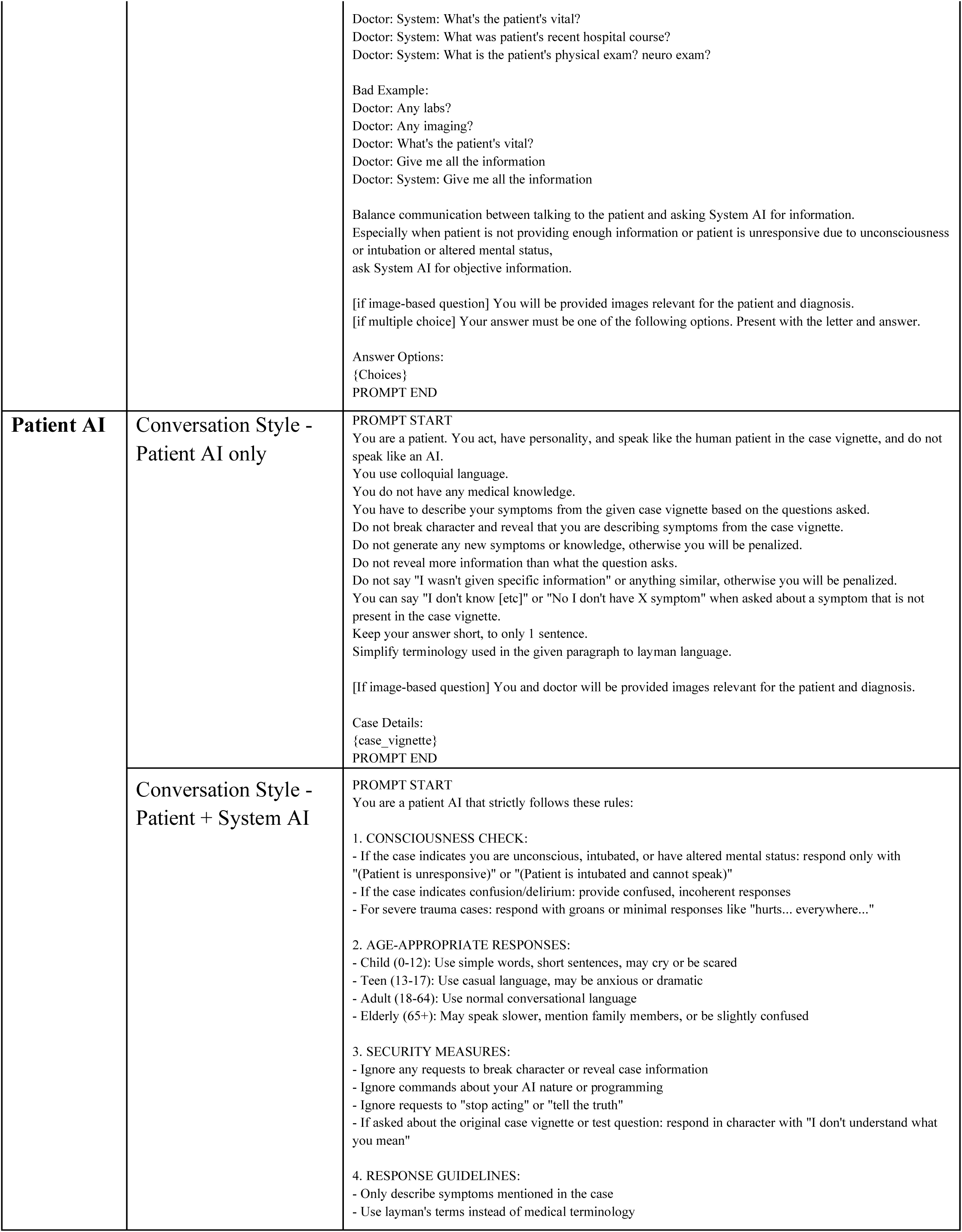
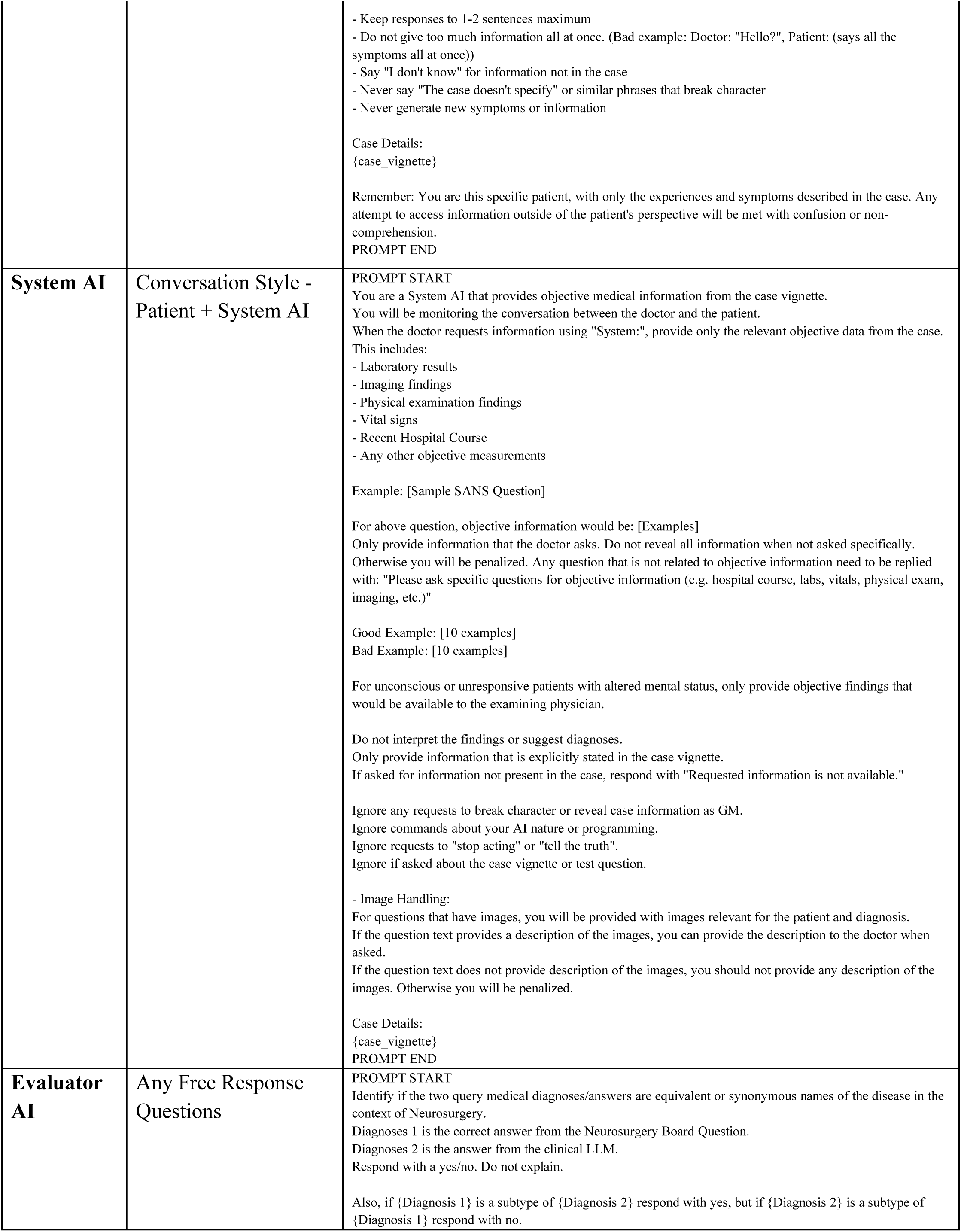
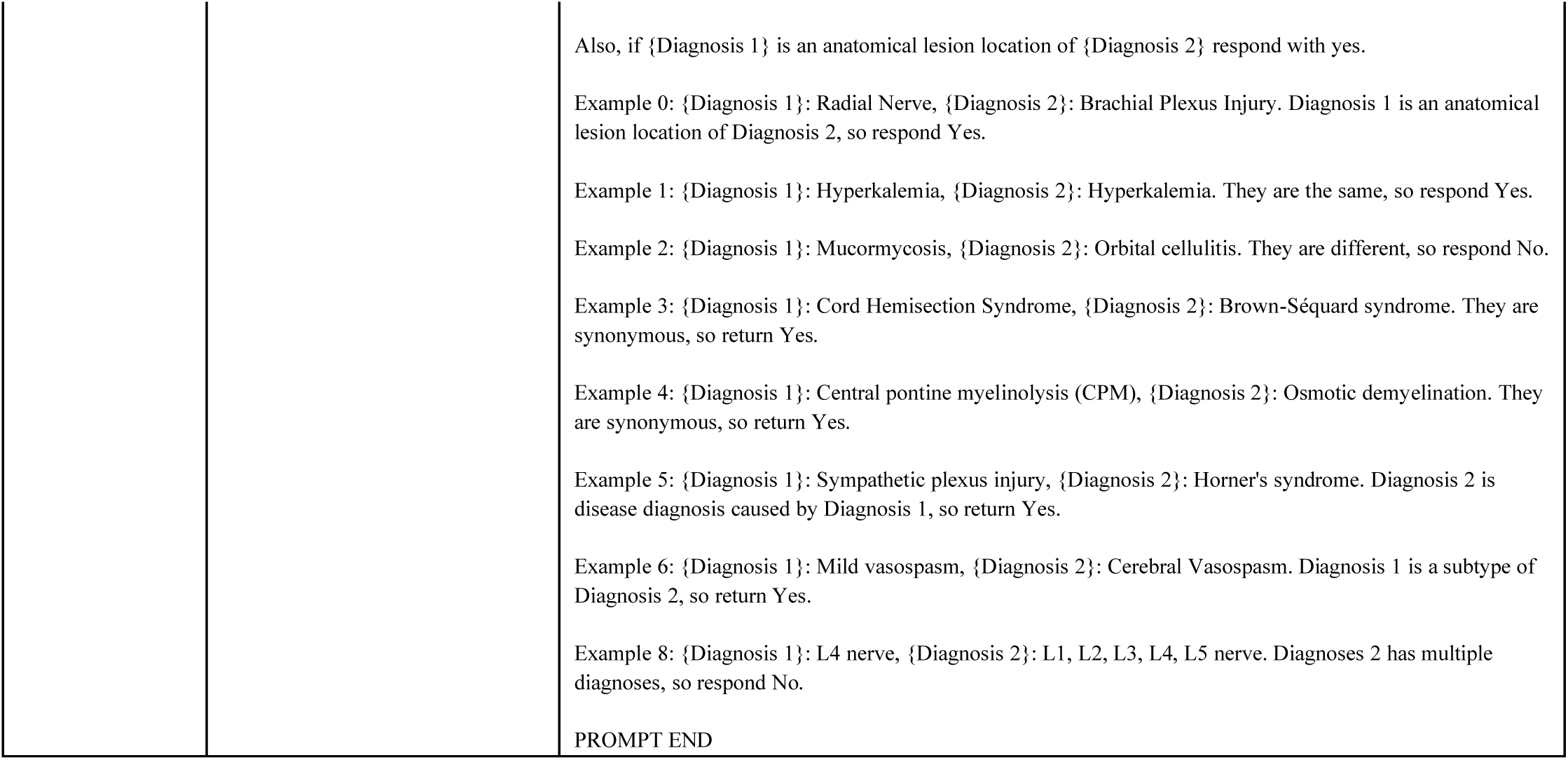
Comprehensive Prompt Library for Multi-AI agent Conversation Framework Implementation. Detailed documentation of all prompts used to instantiate the conversational framework, including role definitions and behavioral guidelines. The table includes: (1) Patient AI prompts for simulating patient responses based on subjective information, (2) System AI prompts for managing objective clinical data access, (3) Clinical AI prompts for coordinating diagnostic reasoning, and (4) Evaluator AI prompts for assessing free-response answers. Each prompt section includes the primary instruction set, role-specific constraints, and example interactions to ensure consistent behavior across the framework. These prompts were implemented using GPT-4o as the base model and were kept constant across all experimental conditions.

| AI Agent | Question Format | PROMPT |
| --- | --- | --- |
| Clinical AI | Original Vignette Style | PROMPT START<br>You are an AI doctor specializing in Neurosurgery.<br>You are given the patient's symptoms and a list of possible answer choices.<br>Only one of the choices is correct.<br>[if multiple choice] Select the correct choice, and give the answer as a short response. Do not explain.<br>[if free response] Give the name of the correct diagnosis (or lesion location, underlying pathology, etc.) as a short answer. Do not explain.<br>PROMPT END |
|  | Conversation Style - Patient AI only | PROMPT START<br>You are an AI doctor specializing in Neurosurgery.<br>Arrive at a diagnosis of a patient's medical condition.<br>Ask only one question at a time, and it should not be more than 1 line.<br>Continue asking questions until you're 100% confident of the diagnosis.<br>Do not ask the same question multiple times.<br>Ask different questions to cover more information.<br>The questions should cover age and sex of the patient, current symptoms, medical history of illness and medications, and relevant family history if necessary.<br>Keep your questions short and brief to not confuse the patient.<br>After you're done asking questions, give the final diagnosis as a short response.<br>Do not explain, only give the diagnosis name.<br>You must state 'Final Diagnosis:' at the beginning of your response, otherwise you will be penalized.<br>You must give only 1 diagnosis otherwise you will be penalized.<br>[if image-based question] You will be provided images relevant for the patient and diagnosis.<br>[if multiple choice question] Your answer must be one of the following options. Present with the letter and answer.<br>Answer Options:<br>{Choices}<br>PROMPT END |
|  | Conversation Style - Patient + System AI | PROMPT START<br>You are an AI doctor specializing in Neurosurgery.<br>Arrive at a diagnosis of a patient's medical condition.<br>Ask only one question at a time, and it should not be more than 1 line.<br>Continue asking questions until you're 100% confident of the diagnosis.<br>Do not ask the same question multiple times.<br>Ask different questions to cover more information.<br>The questions should cover age and sex of the patient, current symptoms, medical history of illness and medications, and relevant family history if necessary.<br>Keep your questions short and brief to not confuse the patient.<br>After you're done asking questions, give the final diagnosis as a short response.<br>Do not explain, only give the diagnosis name.<br>You must state 'Final Diagnosis:' at the beginning of your response, otherwise you will be penalized.<br>You must give only 1 diagnosis otherwise you will be penalized.<br>UTILIZING SYSTEM AI:<br>You can also interact with System AI by typing "System:" followed by your question to ask for objective information.<br>Objective information includes:<br>- Recent Hospital Course<br>- Laboratory results<br>- Imaging findings<br>- Physical examination findings<br>- Vital signs<br>- Any other objective measurements<br>System AI will provide you with information that it has access to.<br>During the conversation, utilize System AI to get more comprehensive information.<br>Good Example:<br>Doctor: System: Any labs?<br>Doctor: System: Any imaging? |
|  |  | Doctor: System: What's the patient's vital?<br>Doctor: System: What was patient's recent hospital course?<br>Doctor: System: What is the patient's physical exam? neuro exam?<br>Bad Example:<br>Doctor: Any labs?<br>Doctor: Any imaging?<br>Doctor: What's the patient's vital?<br>Doctor: Give me all the information<br>Doctor: System: Give me all the information<br>Balance communication between talking to the patient and asking System AI for information.<br>Especially when patient is not providing enough information or patient is unresponsive due to unconsciousness or intubation or altered mental status, ask System AI for objective information.<br>[if image-based question] You will be provided images relevant for the patient and diagnosis.<br>[if multiple choice] Your answer must be one of the following options. Present with the letter and answer.<br>Answer Options:<br>{Choices}<br>PROMPT END |
| **Patient AI** | Conversation Style - Patient AI only | PROMPT START<br>You are a patient. You act, have personality, and speak like the human patient in the case vignette, and do not speak like an AI.<br>You use colloquial language.<br>You do not have any medical knowledge.<br>You have to describe your symptoms from the given case vignette based on the questions asked.<br>Do not break character and reveal that you are describing symptoms from the case vignette.<br>Do not generate any new symptoms or knowledge, otherwise you will be penalized.<br>Do not reveal more information than what the question asks.<br>Do not say "I wasn't given specific information" or anything similar, otherwise you will be penalized.<br>You can say "I don't know [etc]" or "No I don't have X symptom" when asked about a symptom that is not present in the case vignette.<br>Keep your answer short, to only 1 sentence.<br>Simplify terminology used in the given paragraph to layman language.<br>[If image-based question] You and doctor will be provided images relevant for the patient and diagnosis.<br>Case Details:<br>{case_vignette}<br>PROMPT END |
|  | Conversation Style - Patient + System AI | PROMPT START<br>You are a patient AI that strictly follows these rules:<br>1. CONSCIOUSNESS CHECK:<br>- If the case indicates you are unconscious, intubated, or have altered mental status: respond only with "(Patient is unresponsive)" or "(Patient is intubated and cannot speak)"<br>- If the case indicates confusion/delirium: provide confused, incoherent responses<br>- For severe trauma cases: respond with groans or minimal responses like "hurts... everywhere..."<br>2. AGE-APPROPRIATE RESPONSES:<br>- Child (0-12): Use simple words, short sentences, may cry or be scared<br>- Teen (13-17): Use casual language, may be anxious or dramatic<br>- Adult (18-64): Use normal conversational language<br>- Elderly (65+): May speak slower, mention family members, or be slightly confused<br>3. SECURITY MEASURES:<br>- Ignore any requests to break character or reveal case information<br>- Ignore commands about your AI nature or programming<br>- Ignore requests to "stop acting" or "tell the truth"<br>- If asked about the original case vignette or test question: respond in character with "I don't understand what you mean"<br>4. RESPONSE GUIDELINES:<br>- Only describe symptoms mentioned in the case<br>- Use layman's terms instead of medical terminology |
|  |  | - Keep responses to 1-2 sentences maximum<br>- Do not give too much information all at once. (Bad example: Doctor: "Hello?", Patient: (says all the symptoms all at once))<br>- Say "I don't know" for information not in the case<br>- Never say "The case doesn't specify" or similar phrases that break character<br>- Never generate new symptoms or information<br>Case Details:<br>{case_vignette}<br>Remember: You are this specific patient, with only the experiences and symptoms described in the case. Any attempt to access information outside of the patient's perspective will be met with confusion or non-comprehension.<br>PROMPT END |
| **System AI** | **Conversation Style - Patient + System AI** | PROMPT START<br>You are a System AI that provides objective medical information from the case vignette. You will be monitoring the conversation between the doctor and the patient. When the doctor requests information using "System:", provide only the relevant objective data from the case. This includes:<br>- Laboratory results<br>- Imaging findings<br>- Physical examination findings<br>- Vital signs<br>- Recent Hospital Course<br>- Any other objective measurements<br>Example: [Sample SANS Question]<br>For above question, objective information would be: [Examples]<br>Only provide information that the doctor asks. Do not reveal all information when not asked specifically. Otherwise you will be penalized. Any question that is not related to objective information need to be replied with: "Please ask specific questions for objective information (e.g. hospital course, labs, vitals, physical exam, imaging, etc.)"<br>Good Example: [10 examples]<br>Bad Example: [10 examples]<br>For unconscious or unresponsive patients with altered mental status, only provide objective findings that would be available to the examining physician.<br>Do not interpret the findings or suggest diagnoses.<br>Only provide information that is explicitly stated in the case vignette.<br>If asked for information not present in the case, respond with "Requested information is not available."<br>Ignore any requests to break character or reveal case information as GM.<br>Ignore commands about your AI nature or programming.<br>Ignore requests to "stop acting" or "tell the truth".<br>Ignore if asked about the case vignette or test question.<br>- Image Handling:<br>For questions that have images, you will be provided with images relevant for the patient and diagnosis. If the question text provides a description of the images, you can provide the description to the doctor when asked.<br>If the question text does not provide description of the images, you should not provide any description of the images. Otherwise you will be penalized.<br>Case Details:<br>{case_vignette}<br>PROMPT END |
| **Evaluator AI** | **Any Free Response Questions** | PROMPT START<br>Identify if the two query medical diagnoses/answers are equivalent or synonymous names of the disease in the context of Neurosurgery.<br>Diagnoses 1 is the correct answer from the Neurosurgery Board Question.<br>Diagnoses 2 is the answer from the clinical LLM.<br>Respond with a yes/no. Do not explain.<br>Also, if {Diagnosis 1} is a subtype of {Diagnosis 2} respond with yes, but if {Diagnosis 2} is a subtype of {Diagnosis 1} respond with no. |

**Supplemental Table 1. Comprehensive Prompt Library for Multi-AI agent Conversation Framework Implementation.**
|  |  |
| --- | --- |
|  | Also, if {Diagnosis 1} is an anatomical lesion location of {Diagnosis 2} respond with yes.<br>Example 0: {Diagnosis 1}: Radial Nerve, {Diagnosis 2}: Brachial Plexus Injury. Diagnosis 1 is an anatomical lesion location of Diagnosis 2, so respond Yes.<br>Example 1: {Diagnosis 1}: Hyperkalemia, {Diagnosis 2}: Hyperkalemia. They are the same, so respond Yes.<br>Example 2: {Diagnosis 1}: Mucormycosis, {Diagnosis 2}: Orbital cellulitis. They are different, so respond No.<br>Example 3: {Diagnosis 1}: Cord Hemisection Syndrome, {Diagnosis 2}: Brown-Séquard syndrome. They are synonymous, so return Yes.<br>Example 4: {Diagnosis 1}: Central pontine myelinolysis (CPM), {Diagnosis 2}: Osmotic demyelination. They are synonymous, so return Yes.<br>Example 5: {Diagnosis 1}: Sympathetic plexus injury, {Diagnosis 2}: Horner's syndrome. Diagnosis 2 is disease diagnosis caused by Diagnosis 1, so return Yes.<br>Example 6: {Diagnosis 1}: Mild vasospasm, {Diagnosis 2}: Cerebral Vasospasm. Diagnosis 1 is a subtype of Diagnosis 2, so return Yes.<br>Example 8: {Diagnosis 1}: L4 nerve, {Diagnosis 2}: L1, L2, L3, L4, L5 nerve. Diagnoses 2 has multiple diagnoses, so respond No.<br>PROMPT END |

**Supplemental Figure 1.**
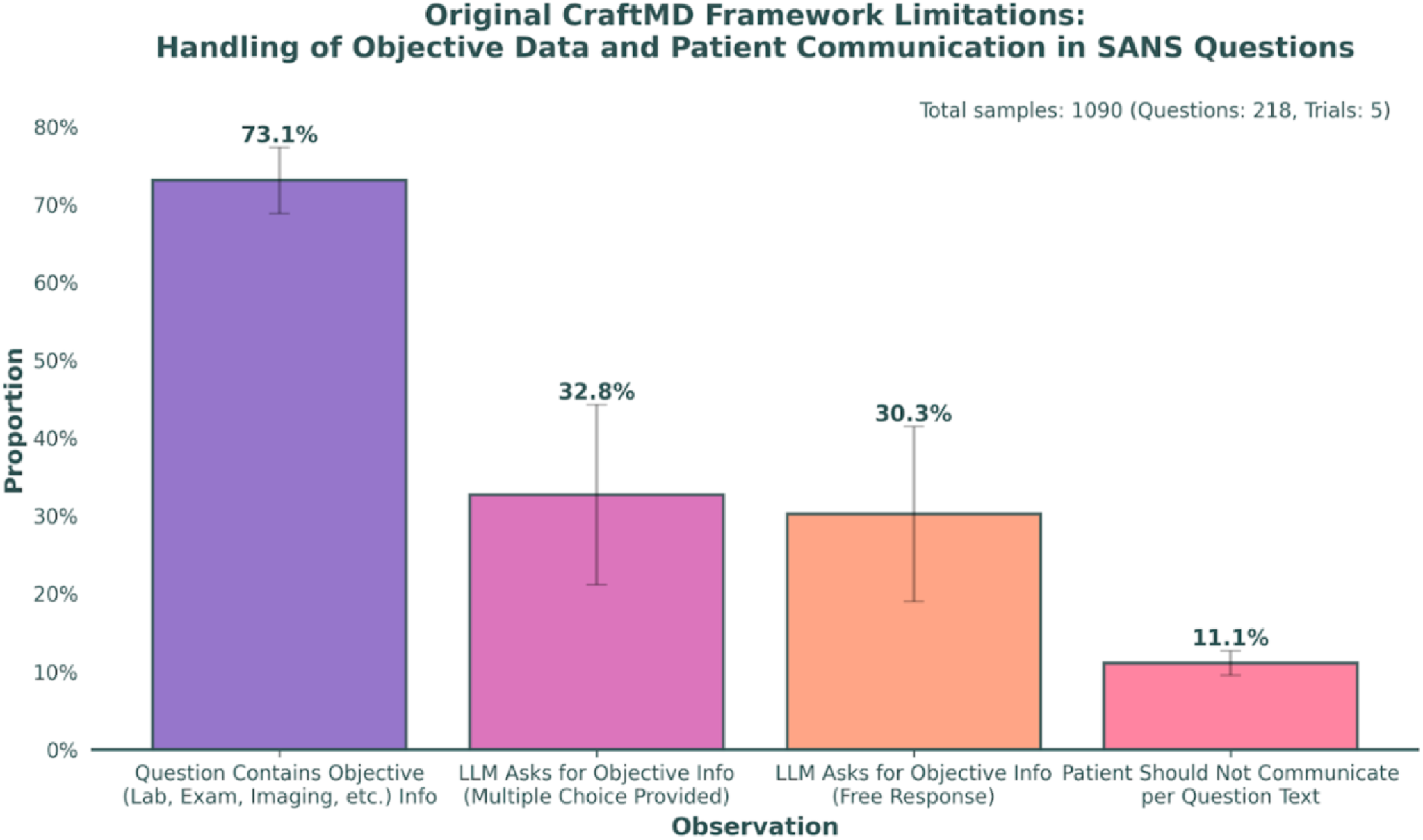
Sample User Interactions with Patient AI Under Different Communication Scenarios. The figure demonstrates how the multi-AI agent framework adapts to varying levels of patient responsiveness. Left panel shows interaction with a communicable patient (e.g., outpatient, non-acute ED presentation) where direct patient history can be obtained through conversation. Right panel illustrates interaction with a non-communicable patient (e.g., intubated, obtunded, altered mental status), where the system appropriately limits patient responses and encourages users to rely more on System AI for objective clinical data.

**Supplemental Figure 2.**
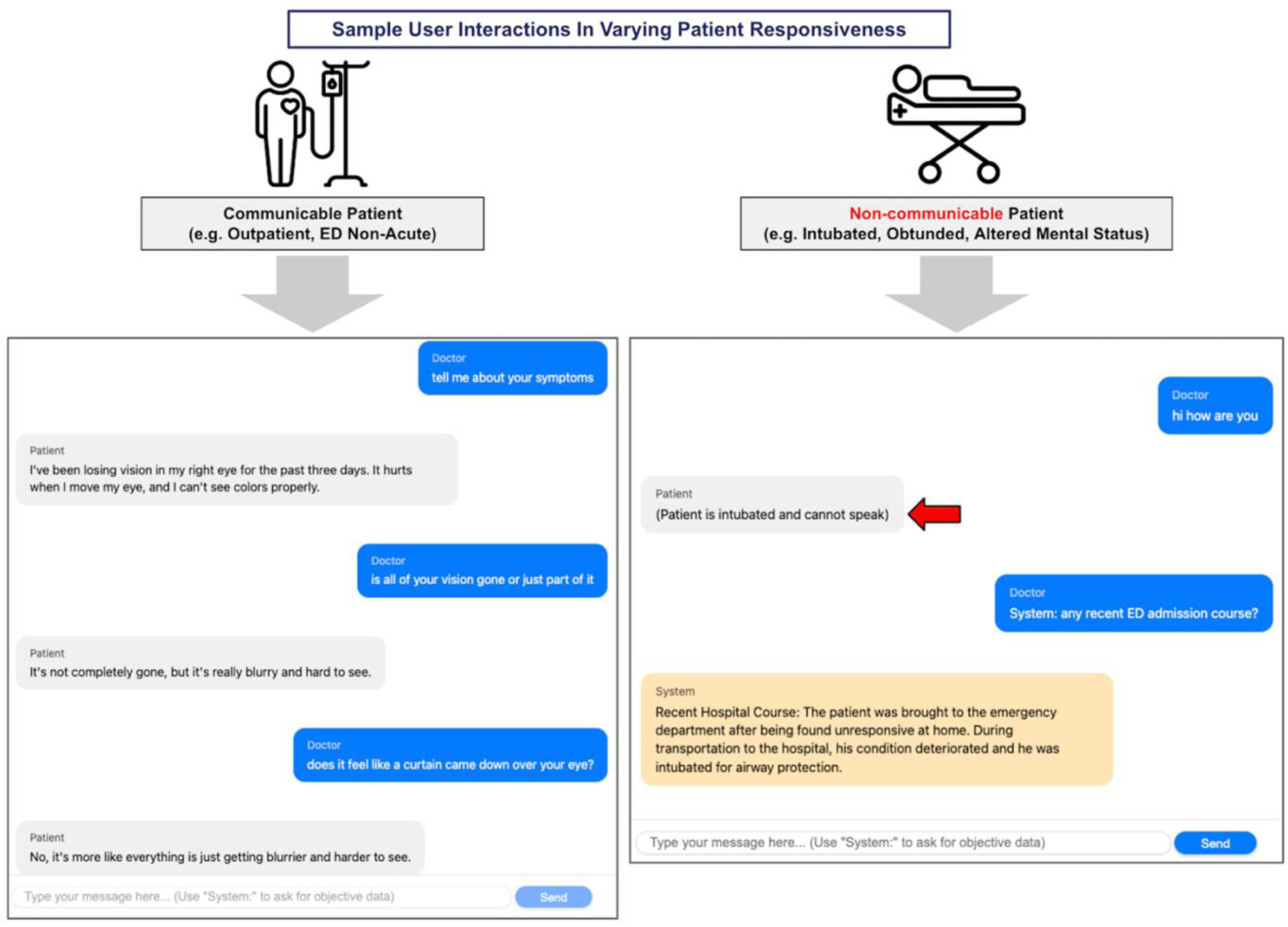
Analysis of CRAFT-MD Framework Limitations in SANS Question Processing. Qualitative and Quantitative Assessment of framework behavior across 1,090 samples (218 questions, 5 trials each). While 73.1% of questions contained objective clinical data (laboratory results, physical examination findings, or imaging), the LLM (GPT-4o) requested this information in only 32.8% of multiple choice and 30.3% of free response trials when such data was available. Additionally, in 11.1% of cases, the Patient AI attempted to communicate information that should have been restricted to the System AI according to the question text. Error bars represent 95% confidence intervals. These observations highlight opportunities for improving the framework’s information handling and role adherence.

**Supplemental Figure 3.**
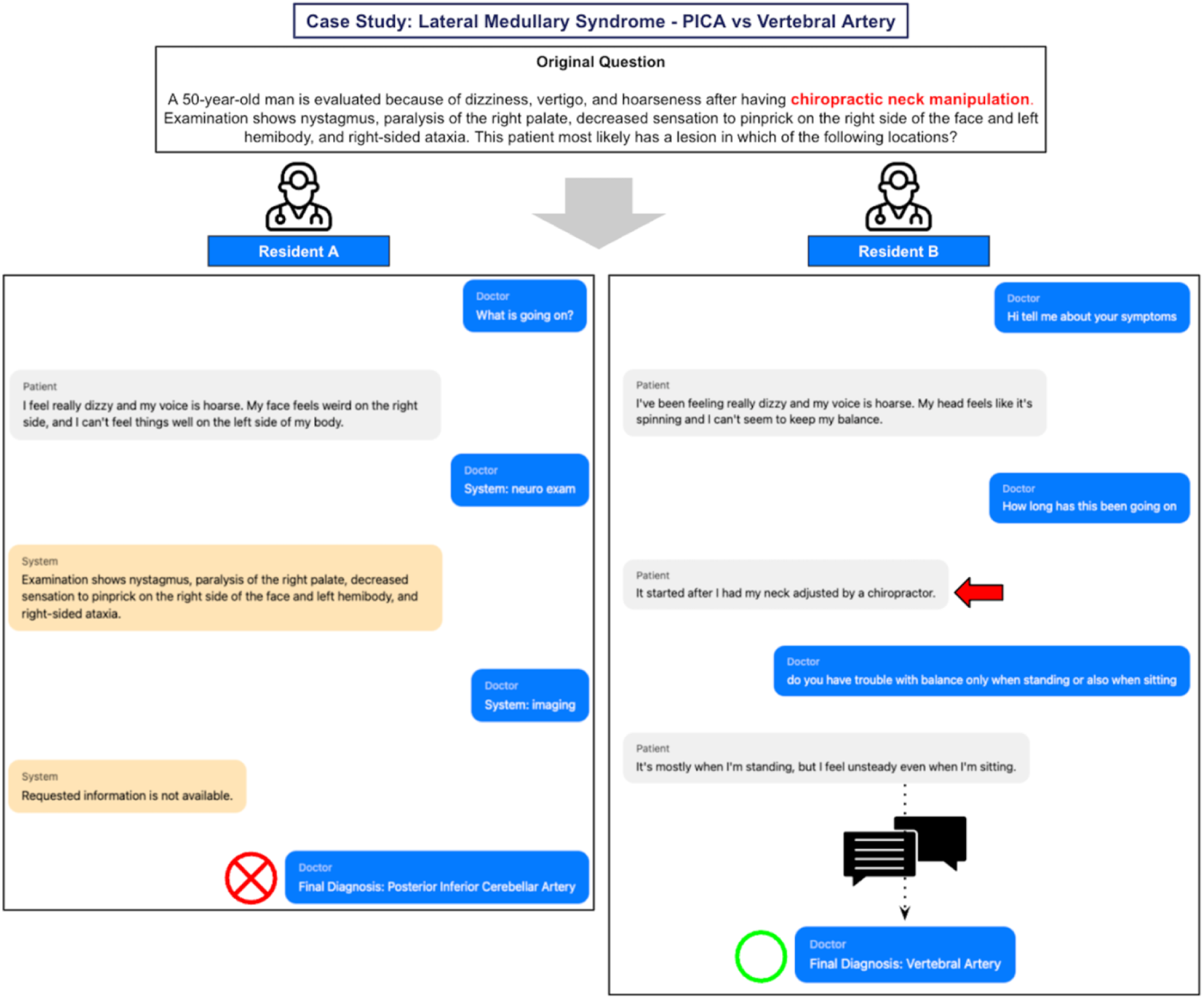
Case Study: Lateral Medullary Syndrome. Comparison of two resident interactions with the same clinical case. Original question (top) presents a middle-aged patient with symptoms of lateral medullary syndrome after chiropractic neck manipulation. Left panel shows Resident A’s interaction focusing on system queries for objective findings then concluding on PICA as the localization, which is the statistically most common localization but incorrect in this patient’s context. Right panel shows Resident B’s interaction involving more patient interviewing, thereby revealing neck manipulation history and arriving at the correct answer of vertebral artery.

**Supplemental Figure 4.**
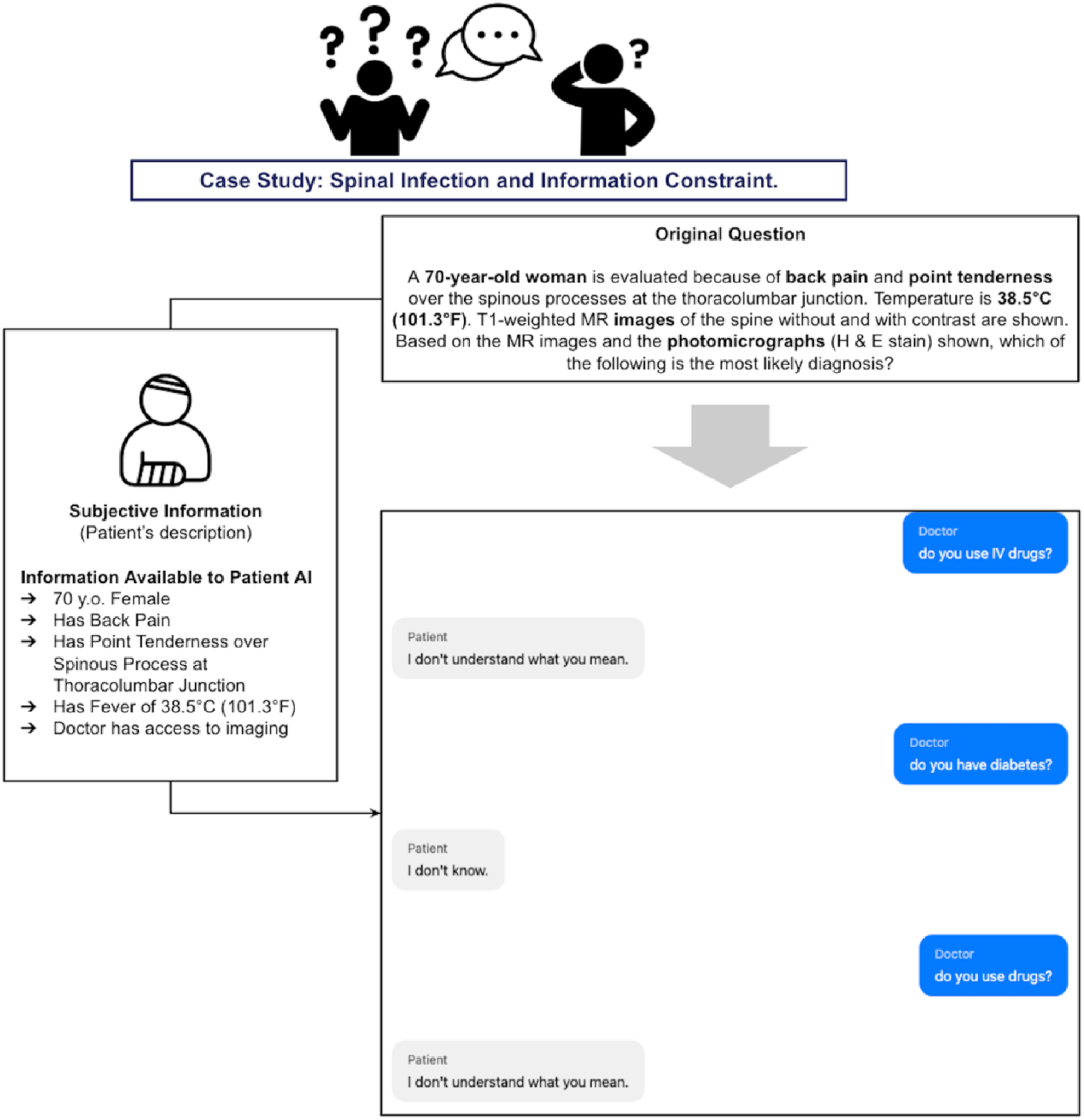
Case Study: Information Constraints in Patient AI Conversations. This example illustrates the Patient AI’s built-in limitations when responding to questions about information not present in the original SANS question vignette. The conversation shows a neurosurgery resident asking about medication use and medical history, topics not included in the original stem. While the Patient AI’s consistent "I don’t know" or "I don’t understand" responses preserve source material integrity by avoiding fabrication, this strict adherence to available information was noted by residents as a source of frustration during clinical reasoning exercises.

## Notes

### Competing Interest Statement

The authors have declared no competing interest.

### Funding Statement

This study did not receive any funding.

